# Genomic determination of relative risks for *Clostridioides difficile* infection from asymptomatic carriage in ICU patients

**DOI:** 10.1101/2020.01.29.20019489

**Authors:** Jay Worley, Mary L. Delaney, Christopher K. Cummins, Andrea DuBois, Michael Klompas, Lynn Bry

**Author notes:** Corresponding author, Lynn Bry, MD, PhD.

## Abstract

**Background:** *Clostridioides difficile* infections (CDIs) are among the most prevalent hospital-associated infections (HAIs), particularly for intensive care unit (ICU) patients. The risks for developing active CDI from asymptomatic carriage of *C. difficile* are not well understood.

**Methods:** We identified asymptomatic *C. difficile* carriage among 1897 ICU patients, using rectal swabs from an existing ICU vancomycin-resistant *Enterococci* (VRE) surveillance program. *C. difficile* isolates from VRE swabs, and from *C. difficile*-positive stool samples, were genome sequenced to assess clonal relationships among isolates from asymptomatic carriers and CDI patients. Integrated genomic and epidemiologic analyses identified multiple cases of asymptomatic carriers who developed CDI, and of asymptomatic transmission of *C. difficile* to naïve patients.

**Results:** Genomic analyses identified diverse strains in infected patients and asymptomatic carriers. 7.4% of ICU patients asymptomatically carried *C. difficile*. 69% of isolates carried an intact toxin locus. In contrast, 96% of *C. difficile* stool isolates were toxigenic. CDI rates in asymptomatic carriers of toxigenic strains were 5.3%, versus 0.57% in non-carriers. The relative risk for CDI with asymptomatic carriage of a toxigenic strain was 9.32 (95% CI=3.25-26.7). Genomic identification of clonal clusters supported epidemiologic analyses for asymptomatic transmission events, with spatial-temporal overlaps identified in 13 of 28 cases.

**Conclusions:** Our studies provide the first genomically-confirmed assessments of CDI relative risk from asymptomatic carriage of toxigenic strains and highlight the complex dynamics of asymptomatic transmission in ICUs. *C. difficile* screening can be implemented within existing HAI surveillance programs and, with isolation of asymptomatic carriers, has potential to reduce these risks.

**Summary:** Relative risks for *C. difficile* infections rise to 9.32 in asymptomatic ICU patients carrying toxigenic strains. Integrated genomic and epidemiologic analyses illustrate functional use of *C. difficile* genomic data to identify asymptomatic transmission events and assist in outbreak investigations.

## Introduction

*Clostridioides difficile* infection (CDI) is the most prevalent healthcare-associated pathogen [1], costing >$5 billion annually in the US, from >500,000 infections and >29,000 deaths [2]. Risks for CDI include chronic contact with healthcare systems, use of broad-spectrum antibiotics, and underlying medical conditions including inflammatory bowel disease or prior CDI [3]. Asymptomatic carriage has also been linked to increased risks for CDI [4]. CDI occurs from pathogen-released toxins, particularly toxins A and B encoded by its pathogenicity locus [5]. A third toxin, binary toxin (CDT), is associated with more severe disease [6].

Asymptomatic carriers are thought to contribute to pathogen spread in healthcare facilities [7–11], but their contributions to underlying pathogen reservoirs, and to their own risks for CDI, are poorly defined [12–14]. One interventional study that placed asymptomatic carriers on contact precautions successfully decreased CDI [15,16]. Healthcare workers have also been identified as potential carriers of toxigenic strains, but at rates reflective of the general population [17–19]. However, healthcare workers can transmit *C. difficile* among patients if standard infection control practices are not followed [20].

We undertook ICU surveillance for *C. difficile* over an 8-month period. Analyses validated sensitivity of a culture-based screening method for *C. difficile* using rectal swabs collected for an existing ICU screening program for vancomycin resistant enterococci (VRE) [4,13,21,22]. Integrated genomic and epidemiologic analyses identified strain dynamics and supported development of a platform leveraging clinical infrastructure and nationally available resources to improve surveillance efforts for *C. difficile* [23].

## Materials and Methods

### IRB Study Protocol and Data Collection

The study was carried out under IRB protocol 2011-P-002883 (Partners Healthcare). The Crimson LIMS [24] was used for sample retrieval over the 246 day study period. VRE-swab retrieval occurred from days 48-199 from medical, surgical, and neurological ICUs (Figure 1), and *C. difficile* toxin-B positive stool collection over the entire study period. Patient demographics and hospital contact data were retrieved from the Partners Research Patient Data Registry and Theradoc [25], and were de-identified for analyses (Table 1; Supplemental Table 1).

**Table 1:**
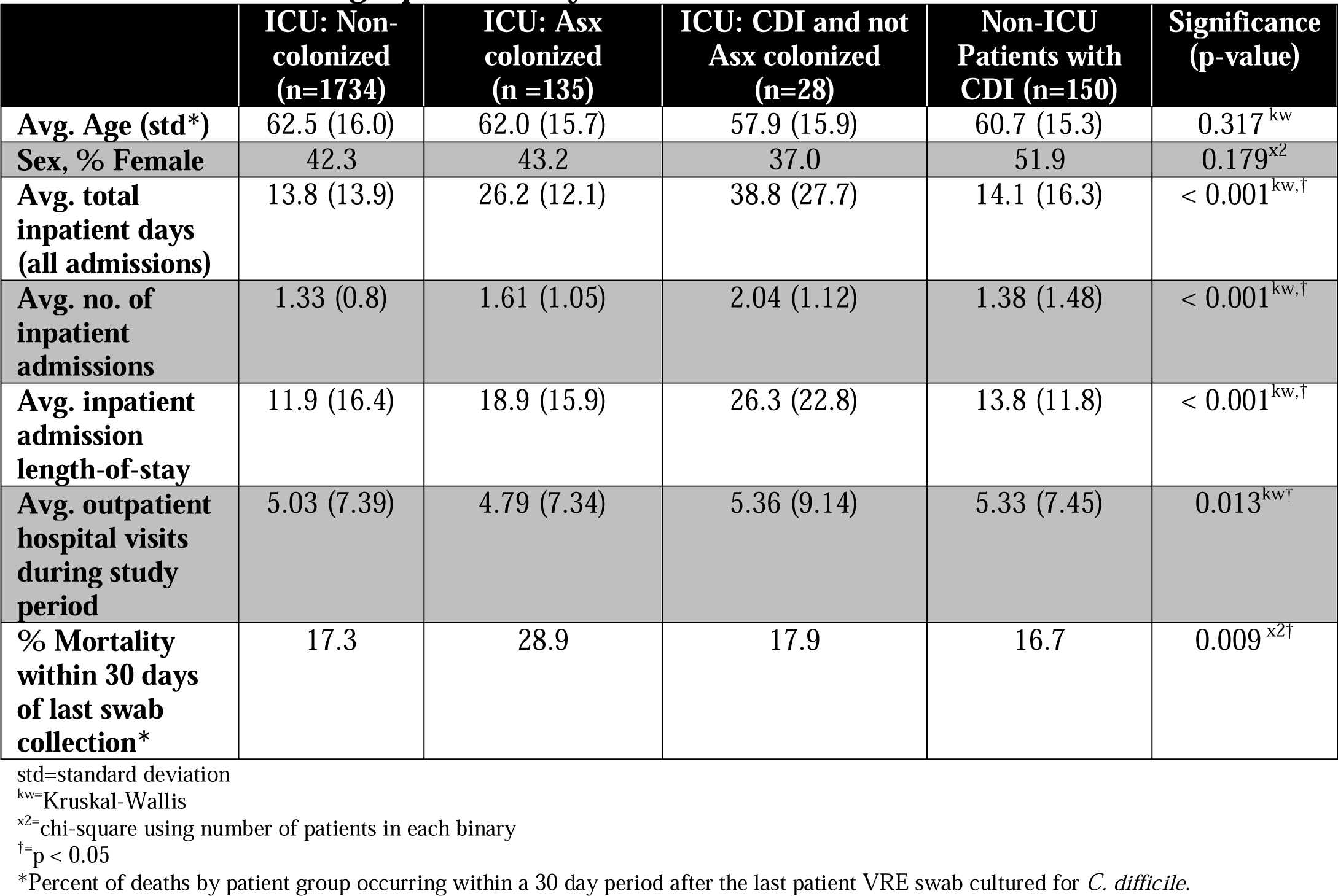
Patient demographic data by *C. difficile* colonization and CDI status.

|  | ICU: Non-colonized<br>(n=1734) | ICU: Asx colonized<br>(n =135) | ICU: CDI and not<br>Asx colonized<br>(n=28) | Non-ICU<br>Patients with<br>CDI (n=150) | Significance<br>(p-value) |
| --- | --- | --- | --- | --- | --- |
| Avg. Age (std*) | 62.5 (16.0) | 62.0 (15.7) | 57.9 (15.9) | 60.7 (15.3) | 0.317<sup>kw</sup> |
| Sex, % Female | 42.3 | 43.2 | 37.0 | 51.9 | 0.179<sup>x2</sup> |
| Avg. total inpatient days<br>(all admissions) | 13.8 (13.9) | 26.2 (12.1) | 38.8 (27.7) | 14.1 (16.3) | < 0.001<sup>kw,†</sup> |
| Avg. no. of inpatient admissions | 1.33 (0.8) | 1.61 (1.05) | 2.04 (1.12) | 1.38 (1.48) | < 0.001<sup>kw,†</sup> |
| Avg. inpatient admission length-of-stay | 11.9 (16.4) | 18.9 (15.9) | 26.3 (22.8) | 13.8 (11.8) | < 0.001<sup>kw,†</sup> |
| Avg. outpatient hospital visits during study period | 5.03 (7.39) | 4.79 (7.34) | 5.36 (9.14) | 5.33 (7.45) | 0.013<sup>kw†</sup> |
| % Mortality within 30 days of last swab collection* | 17.3 | 28.9 | 17.9 | 16.7 | 0.009<sup>x2†</sup> |

std=standard deviation
<sup>kw</sup>=Kruskal-Wallis
<sup>x2</sup>= chi-square using number of patients in each binary
<sup>†</sup>= p < 0.05
\*Percent of deaths by patient group occurring within a 30 day period after the last patient VRE swab cultured for *C. difficile*.

**Figure 1:**
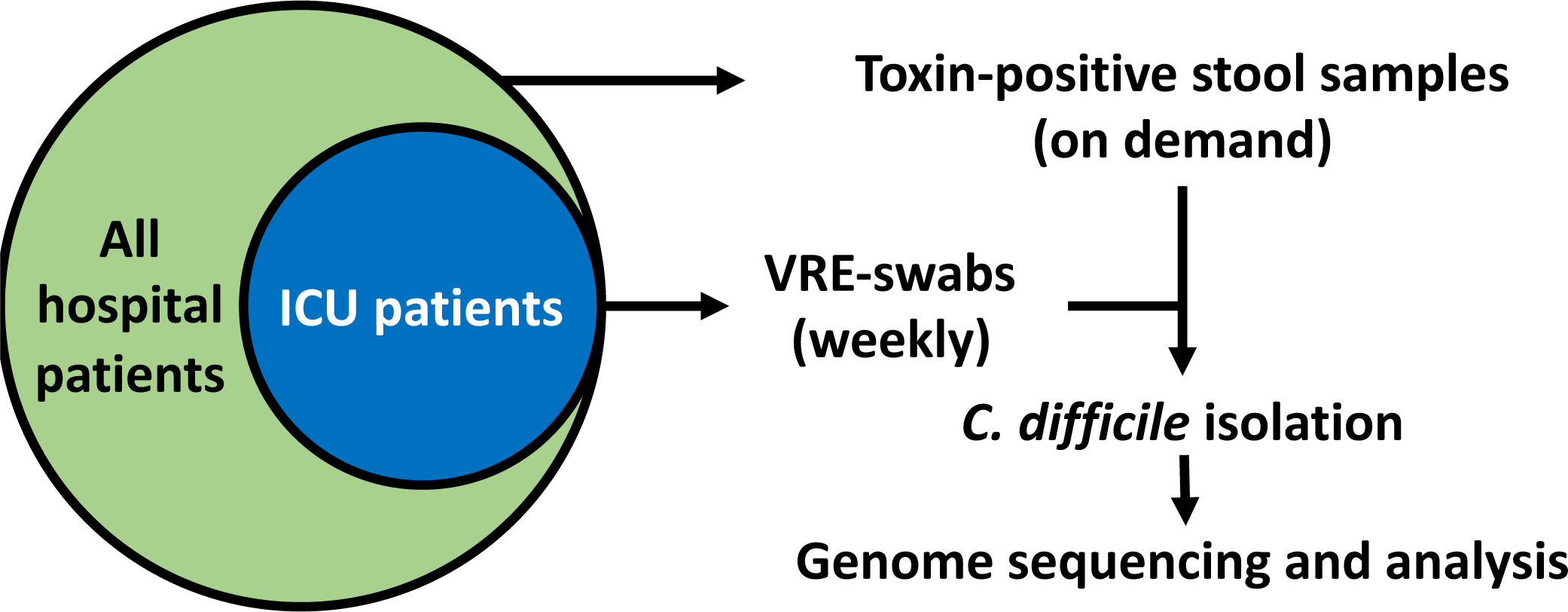
Patient group and sample processing overview. ICU patients are a subset of all patients. *C. difficile* isolation was performed on stool samples that were toxin-positive by EIA, and all VRE-swabs. All isolateswere sequenced.

### Validation of VRE-swabs for detection of *C. difficile* carriage

Two clinical *C. difficile* isolates were grown in Brain Heart Infusion Broth and serially diluted from 1×10^5^-10^2^ CFU/mL. To mimic clinical culture conditions, BD BBL CultureSwabs (BD, San Jose, CA) were inoculated with 100ul of a given dilution, placed in transport tubes and stored aerobically for 2 hours. The swabs were plated to Spectra VRE agar (Thermo Scientific, Waltham, MA) to mimic expected swab analyses for VRE, then returned to their transport tubes and stored for another 2 hours prior to plating on CHROMID *C. difficile* agar (Biomerieux, Durham, NC) with incubation at 37°C in an anaerobic chamber for 24 hours. *C. difficile* colonies were identified as black colonies on CHROMID agar and were confirmed by Gram stain and rapid ANA panels (Biomerieux, Durham, NC).

### Sample collection

BWH ICU patients are screened clinically for VRE by collection of rectal swabs upon ICU admission and weekly thereafter. Clinical stool samples positive for toxin B (TcdB) were retrieved after testing. Samples were plated onto CHROMID *C. difficile* agar and confirmed isolates of *C. difficile* subjected to genomic analyses.

### Genomic analyses

Genomic sequencing by Illumina MiSeq was performed as described (Illumina, San Diego, CA) [24]. Genome assembly was done using SPAdes [26] (Supplemental File 1). Toxin typing used reference *tcd* toxin genes from CD630 (AM180355.1). *cdt* toxin reference gene content used strain R20291 (NC_013316.1). Gene calling used cutoffs of 80% reference gene length and 80% amino acid sequence identity. SNP-based analyses used the NCBI Pathogen Detection Isolates Browser (https://www.ncbi.nlm.nih.gov/pathogens/) [23].

### *C. difficile* Phylogenetic Analyses

*C. difficile* genomes from the Sequence Reads Archive (SRA) (https://ncbi.nlm.nih.gov/SRA) and NCBI Pathogen Detection Isolates Browser (https://ncbi.nlm.nih.gov/pathogens/isolates) were downloaded for analyses. SPAdes draft assembled genomes passing the following criteria were used: genome sequence length within 3.7-5.0 Mb and <150 contigs, an L90 (fewest number of contigs that cover 90% of assembled sequence) >30, or an average coverage >25X. Analyses evaluated 3377 genomes within 173 SNP groups (Supplemental File 2).

A core *C. difficile* genome was created by performing tblastn on the CD630 reference genome. Feature identification used cutoffs of ≥80% protein sequence identity and feature length within 20% of the reference protein sequence length. Nucleotide alignments of the extracted genes used MAFFT [27].

A representative phylogeny within the NCBI Pathogen Detection Isolates Browser was developed using up to three members from each SNP group from a 95% core genome of 2835 genes resulting in a 422,008b SNP matrix. Of these SNP base positions, 353,060 occurred in at least 95% of aligned positions, and were used to calculate a phylogeny in RAxML with 1000 bootstraps and maximum likelihood analyses using the GTRCAT matrix [28]. The clade structure concurred with previous phylogenetic analyses [29], and includes 157 SNP clusters, 256 non-clustered single isolates, and several paraphyletic genomes [30]. The tree is available online at https://itol.embl.de/tree/17022320725465491568233605.

### Statistical analyses

Statistical analyses used the Python package SciPy [31]. Differences in demographic data among patient groups were calculated using the Kruskal-Wallis test and Mannfred-Whitney post-hoc test for continuous variables and χ^2^ test for discreet variables. Multi-hypothesis adjusted p-values were calculated using the Benjamini Hochberg Procedure [32]. Relative risk ratios were calculated by dividing the probability of developing CDI in the asymptomatically colonized (exposed) group of patients by the probability of developing CDI in the control (non-colonized) group (Table 2).

**Table 2:** Relative risk for CDI from asymptomatic carriage of *C. difficile*.

| **Toxin-positive *C. difficile*** | **No CDI** | **CDI** |
| --- | --- | --- |
| VRE-swab, toxigenic *C. difficile* isolate | 89 | 5 |
| VRE-swab, no isolate or non-toxigenic *C. difficile* isolated | 1742 | 10 |
| **Toxin-negative *C. difficile*** | **No CDI** | **CDI** |
| VRE-swab, toxigenic *C. difficile* isolate | 45 | 0 |
| VRE-swab, no isolate or non-toxigenic *C. difficile* isolate | 1786 | 15 |
| ***C. difficile*, toxin positive or negative strain** | **No CDI** | **CDI** |
| VRE-swab, *C. difficile* isolated | 134 | 5 |
| VRE-swab, no isolate | 1697 | 10 |

CDI relative risk from carriage of toxin+ *C. difficile*: 9.32. 95% CI: 3.25 to 26.7, $p < 0.001$
CDI relative risk from carriage of toxin+ or - *C. difficile*: 6.14. 95% CI: 2.13 to 17.7, $p < 0.001$

## Results

### *C. difficile* surveillance program

Brigham & Women’s Hospital in Boston, Massachusetts, is a 793-bed hospital providing care to >600,000 patients/year, including >36,000 inpatient admissions. The hospital includes multiple ICUs. The CDI genomic screening pilot evaluated 2432 VRE-swabs from 1897 ICU patients over 152 days, from which 172 *C. difficile* isolates were identified in 143 patients (Figure 1, Table 1, Supplemental Table 1). Asymptomatic colonization with *C. difficile* occurred in 7.4% (n=140) of patients, including 5 who developed CDI. During this period, and for the 47 days before and after the swab collection period, 177 toxin B-positive stool samples from all hospital patients were cultured, with 98.3% of stool samples (n=174) growing a *C. difficile* isolate. Of these isolates, 28 (16.1%) were from patients with ICU admissions who had VRE-screening performed (Figure 2).

**Figure 2:**
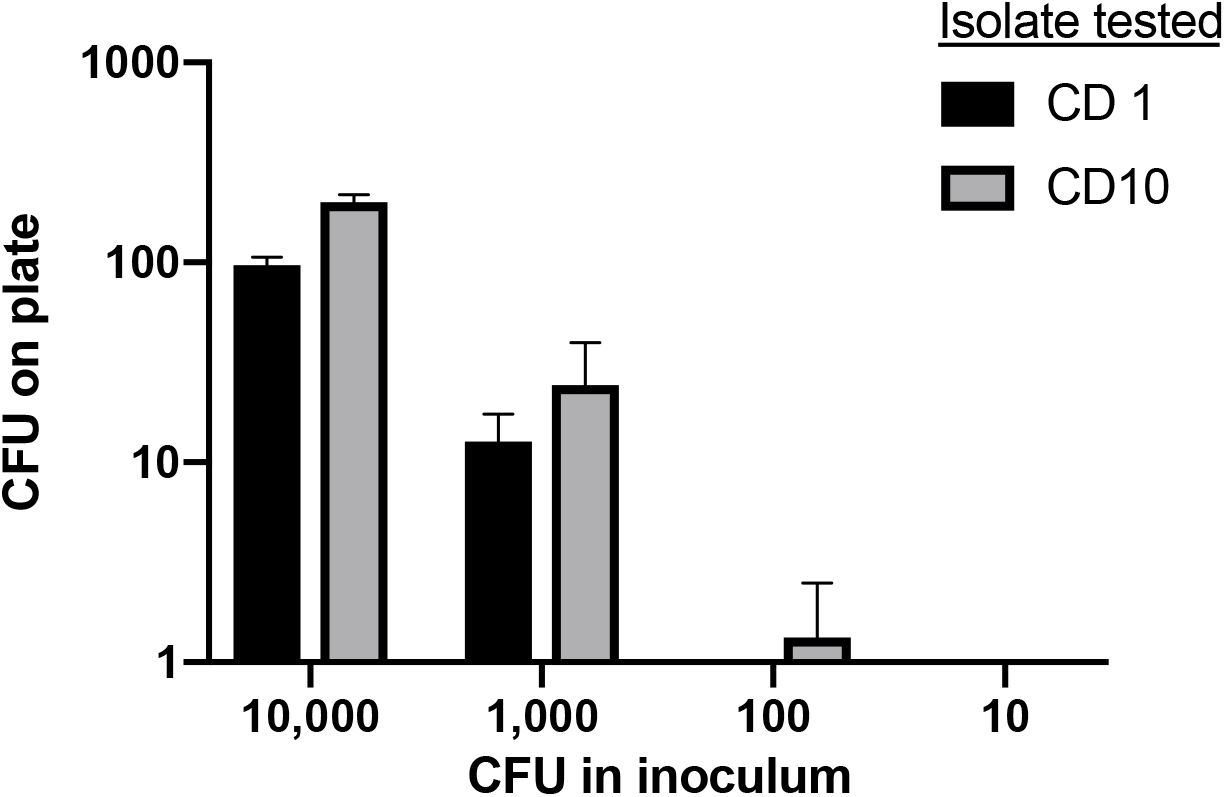
Sensitivity of VRE-swabs with CHROMID agar for *C*. *difficile*. CFU in inoculum were directly applied to swab tips before recapping into Stuart’s media sponge reservoir. CFU on plate are the number of colonies seen after plating onto CHROMID C. difficile agar.

### Association of colonization status with hospital admissions and mortality

Asymptomatic *C. difficile* colonization and CDI development were each associated with increased inpatient days across admissions that were spent in the hospital (Table 1, row 3). ICU patients who developed CDI during the study period averaged 38.8 inpatient days, asymptomatically colonized ICU patients 26.2 days, versus 13.8 days for non-colonized ICU patients (p<0.001). Asymptomatic carriers and CDI-diagnosed ICU patients also had increased lengths-of-stay per hospital admission (Table 1, row 5). ICU patients diagnosed with CDI demonstrated an average of 26.3 days versus 18.9 days for asymptomatic carriers, and 11.9 days per admission for non-colonized ICU patients (p<0.001).

In comparing admission lengths-of-stay between ICU and non-ICU patients, non-ICU patients diagnosed with CDI averaged 13.8 days per admission, whereas ICU and CDI patients averaged 26.3 days per admission (p=0.003).

The differences in length-of-stay among ICU patients were not associated with significant differences in the number of admissions for asymptomatically colonized vs. non-colonized patients, at 1.61 vs. 1.33 admissions during the study period. However, patients with ICU admissions who developed CDI during their hospitalization had higher repeat admissions of 2.04 (p<0.017). Mortality rates, in the 30 days after a patient’s last VRE swab cultured for *C. difficile*, were also higher in asymptomatically colonized versus non-colonized ICU patients at 28.9% vs 17.3%, respectively (p=0.006).

### Genomic diversity of hospital *C. difficile* isolates

For isolate genomic contextualization, genome sequences were submitted to NCBI for comparison with other *C. difficile* genome sequences submitted to NCBI. Strains were placed into SNP clusters by NCBI Pathogen Detection, and these clusters were analyzed for clade designation and ST. The 346 patient isolates were highly diverse and occurred primarily in clades 1 and 2 (Figure 3). Within clade 2, 20 isolates clustered to sequence type 1 (NAP1/RT027, *cdt* toxin positive; 5.8% of isolates). Clade 2 strains were more than twice as likely to originate from CDI stool samples (24 isolates) than from VRE screening swabs (11 isolates; p=0.035). The *cdt* toxin locus was also identified in strains from clade 5, primarily in ST11 from SNP cluster PDS000017348 [33]. CDT-encoding strains occurred more frequently from stool (n=30) than VRE-swabs (n=16; p=0.044).

**Figure 3:**
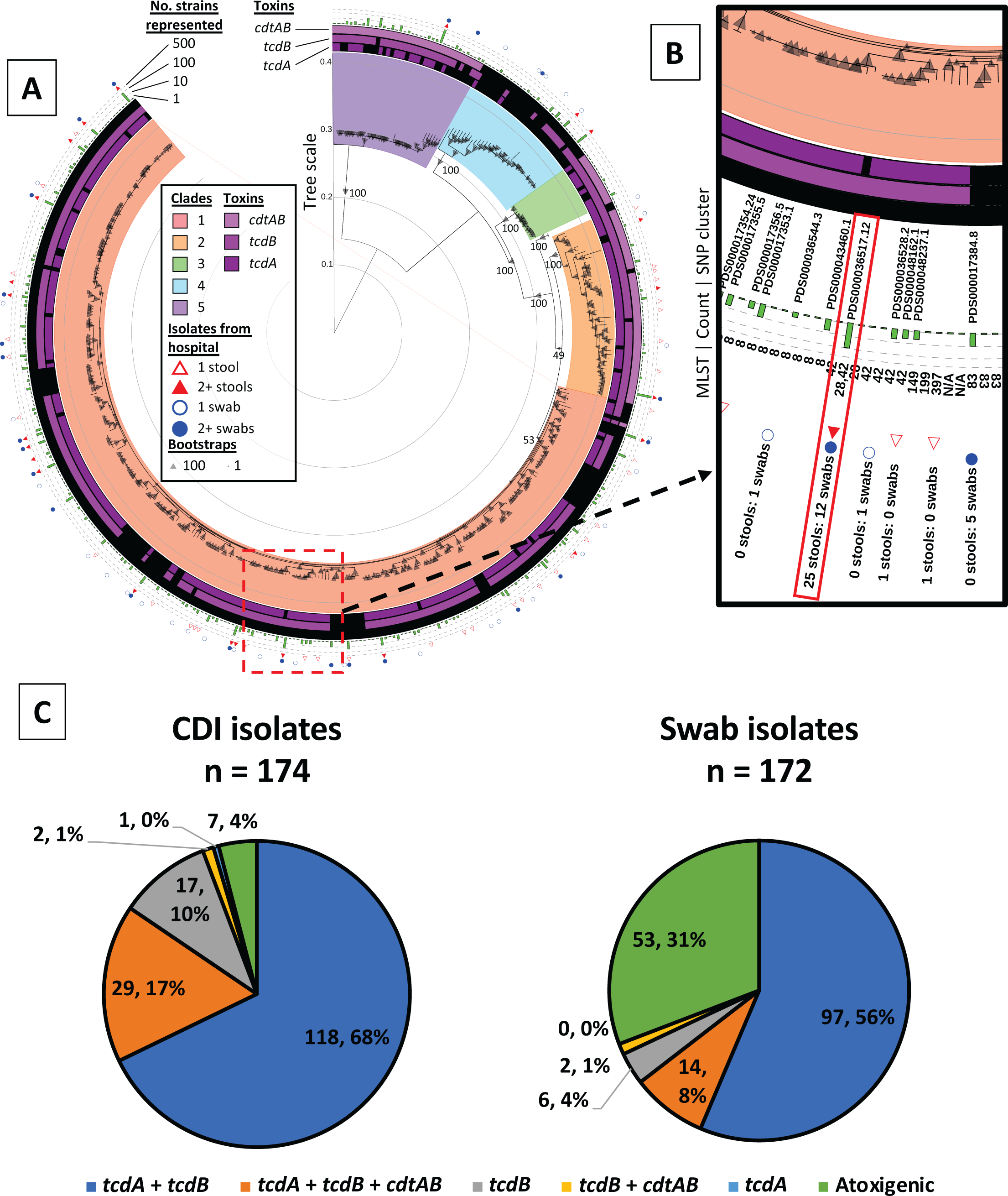
A) Distribution of clinical isolates among all *C. difficile* strains. 157 SNP clusters, each represented by one isolate, and 256 non-clustered isolates are shown. The outer ring shows the type of samples present in the branch, and whether multiple samples were found therein. A bar graph in the second ring shows the number of strains represented by the leaf in the NCBI Pathogen Detection Isolates Browser. The inner rings demonstrate the variability of toxin types found within each clade. Clades are indicated by a colored box around the interior nodes that compose the clade. B) An interactive view available online that shows the sequence types and specifies how many strains are represented in the leaf. C) Percentages of isolates from the different sample types with the indicated encoded toxin phenotypes. The difference in the number of atoxigenic strains is significant at p < 0.001.

Clade 4 included two distinct genetic groups, including ST37 isolates with only toxin B (*tcdB*), of which 7 originated from stool samples and 1 from a VRE-swab, a proportion differing significantly from the remaining atoxigenic Clade 4 strains (n=5; p=0.01).

### Toxin genotypes of strains from CDI patients versus asymptomatic carriers

Among ICU carriers of *C. difficile* 31.8% of strains were atoxigenic lacking both the *tcd* and *cdt* loci (53 isolates from 45 patients). In contrast, only 7 strains (4%) from CDI patients did not encode a detectable toxin locus (p<0.001).

### Longitudinal strain dynamics in asymptomatic ICU carriers

A total of 25 ICU patients (15.3% of all swab-positive patients), including 17 asymptomatic carriers, demonstrated longitudinal *C. difficile* carriage (Figure 4). The longitudinal strain sets were used to evaluate SNP diversity in presumed clonal isolates over time. The maximum number of SNPs was 17, with an average of 7.5 per asymptomatic patient.

**Figure 4:**
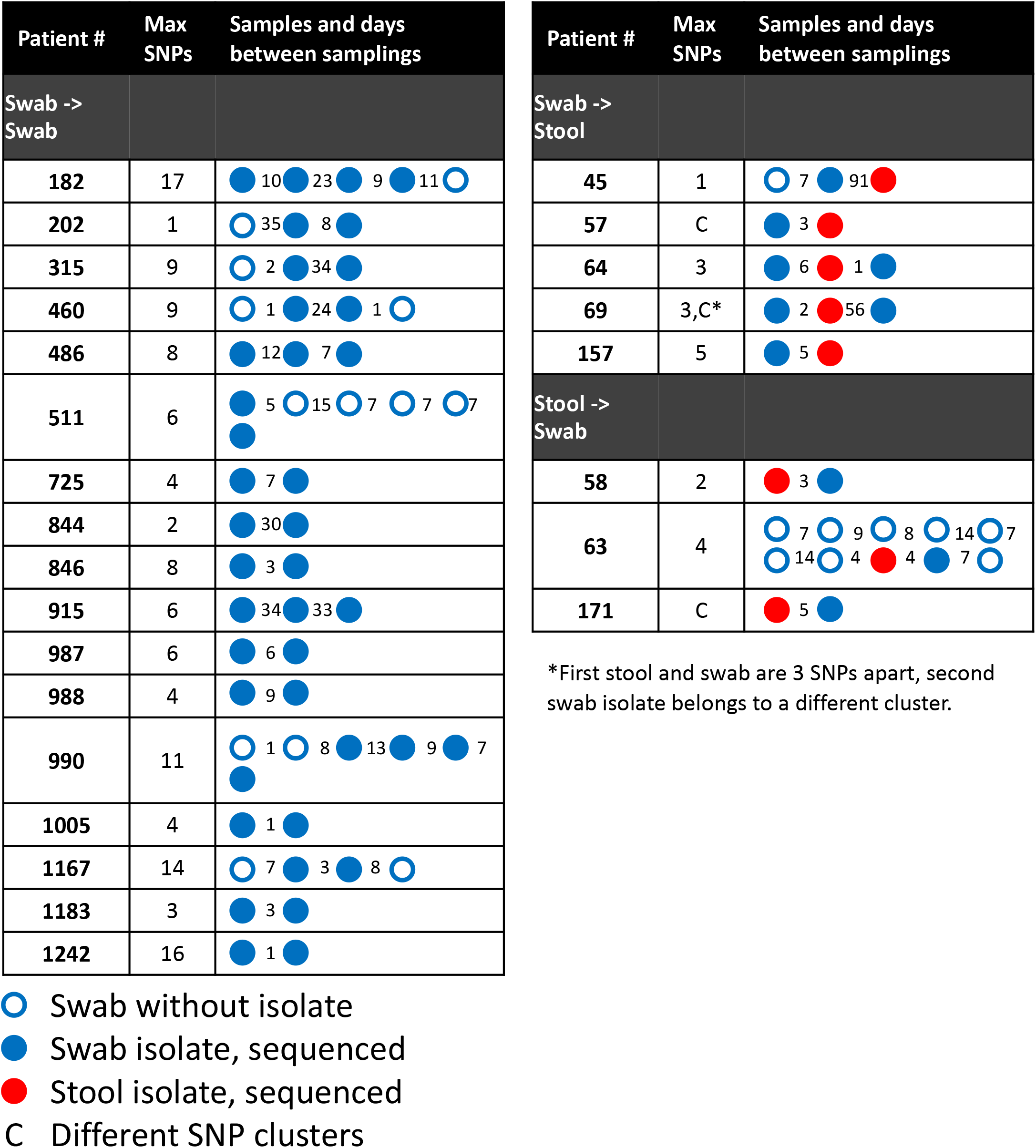
Patients with multiple sequenced isolates. Patient samples are indicated with circles, blue indicating a VRE-swab and red indicating a toxin positive stool. Filled circles are samples that produced isolates that were then sequenced. The number of days between samples is indicated by a number between circles. In all but three cases, all isolates were in the same cluster, and of those three, only in Patient 57 did the strain from a different cluster appear after *C. difficile* diagnosis and the beginning of treatment.

### CDI occurrence in asymptomatic carriers

Five asymptomatic ICU carriers developed active CDI (Figure 4). In four cases, the VRE-swab isolate was within 5 SNPs of the stool isolate and was considered clonally related. These patients developed CDI within 7 days of detecting *C. difficile* from VRE swabs. The 5^th^ case, patient 57, developed CDI with a different epidemic ST37 strain, which occurred 91 days after detection of asymptomatic carriage.

Five separate ICU patients also had *C. difficile*-positive VRE-swabs after CDI diagnosis. In patients 69 and 171, genomic analyses identified that swab isolates from different SNP clusters, suggesting potential acquisition of new strains after clearance from treatment. Patient 63 had a period of 59 days and 7 negative VRE-swabs, followed by CDI onset and continued colonization with the infecting strain before returning to a non-colonized state.

### Relative risks for CDI from asymptomatic carriage of toxigenic *C. difficile*

The relative risk for developing CDI from asymptomatic carriage of a toxigenic strain was 9.32 (95% CI: 3.25-26.7; p<0.001), while carriage of a non-toxigenic strain did not increase risks for CDI; no carriers of non-toxigenic strains developed CDI over the study period (Table 2). The 5 CDI cases from 140 ICU asymptomatic carriers represented 3.6% of carriers. Of the 163 patients who tested positive for *C. difficile* while in an ICU, 82.8% did not develop active CDI for the subsequent 6 months after their last tested swab.

### Genomic-epidemiological investigations of asymptomatic transmission

Genomic cluster analyses identified related sub-clusters of isolates for spatial-temporal analyses to assess potential transmission events (Figure 4). Per the relatedness of longitudinal isolates from the same patient, thresholds of <17 SNPs and individual branch lengths within clusters of <15 SNPs were used to identify clusters for analyses. Analyses identified 28 sub-clusters across 20 SNP groups, involving 76 isolates from 65 patients (Supplemental file 3). Analyses evaluated spatio-temporal linkages among hospital floors, wings within a floor, rooms, and bed spaces within a room.

Within the 28 clusters, the largest sub-cluster of genomically-related strains occurred in SNP cluster PDS000036517 (Figure 5) and included 18 isolates from BWH patients with others reported from the University of Pittsburgh. Isolates within this cluster were 2-34 SNPs apart, ruling out a single outbreak cluster. Three sub-clusters of BWH isolates within this group met criteria for investigation, one of which contained two isolates from the same patient. The other two clusters did not show spatial-temporal overlaps for the affected patients while in the hospital and were ruled out as nosocomial transmission events.

**Figure 5:**
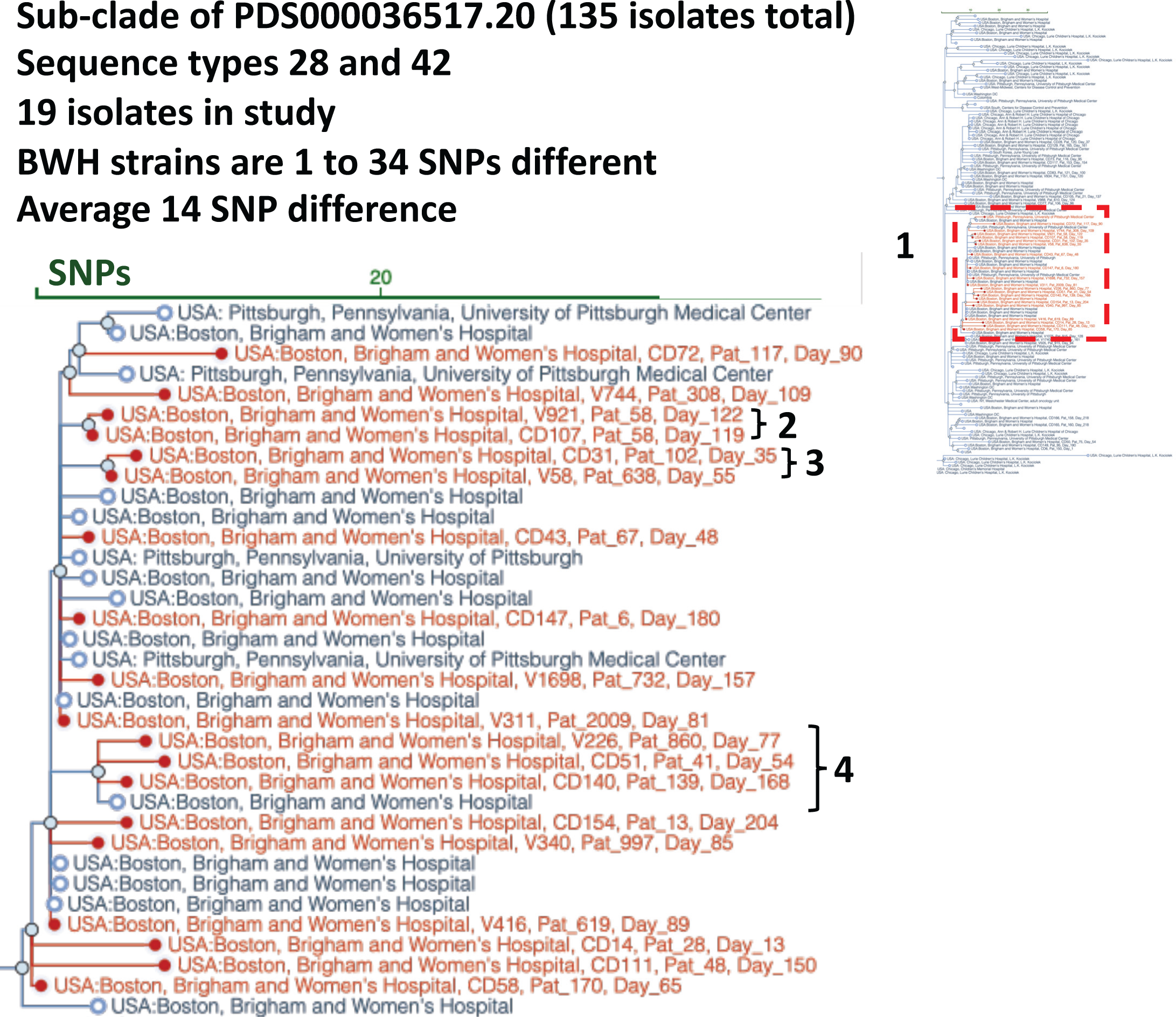
Sub-clade selection. The NCBI Pathogen Detection Isolates Browser provides a comparison tool for identifying outbreaks and relating them to other isolate genomes submitted to NCBI. This subclade of 19 strains is related to others that have been submitted to NCBI from our institution and isolates from other institutions. 1) Overview of the whole SNP cluster. 2) Two isolates from the same patient requiring no further investigation. 3) Two isolates closely related and 20 days apart in isolation. 4) Four isolates forming a related subclade but with one related sample from outside of this study, submitted earlier from Brigham and Women’s Hospital.

Thirteen of 28 clonal clusters had isolates from patients with spatial overlaps within the hospital in the prior 210 days to when the isolate clusters were identified (Figure 6, Supplemental Figure 1). In 26 of the 28 individuals from these overlapping cases, the affected patients shared a floor location, 15 shared a wing location, 3 the same room, and 2 the same bed space. Using a Poisson cumulative probability cutoff of 0.1, an investigative threshold used by local hospital infection control teams to flag clusters for subsequent evaluation, the period where a repeat observation on the same floor could be considered linked is 258 days, and 447 days for observations from the same wing. The 28 cases flagged by initial genomic analyses of related strains fell within these time constraints.

**Figure 6:**
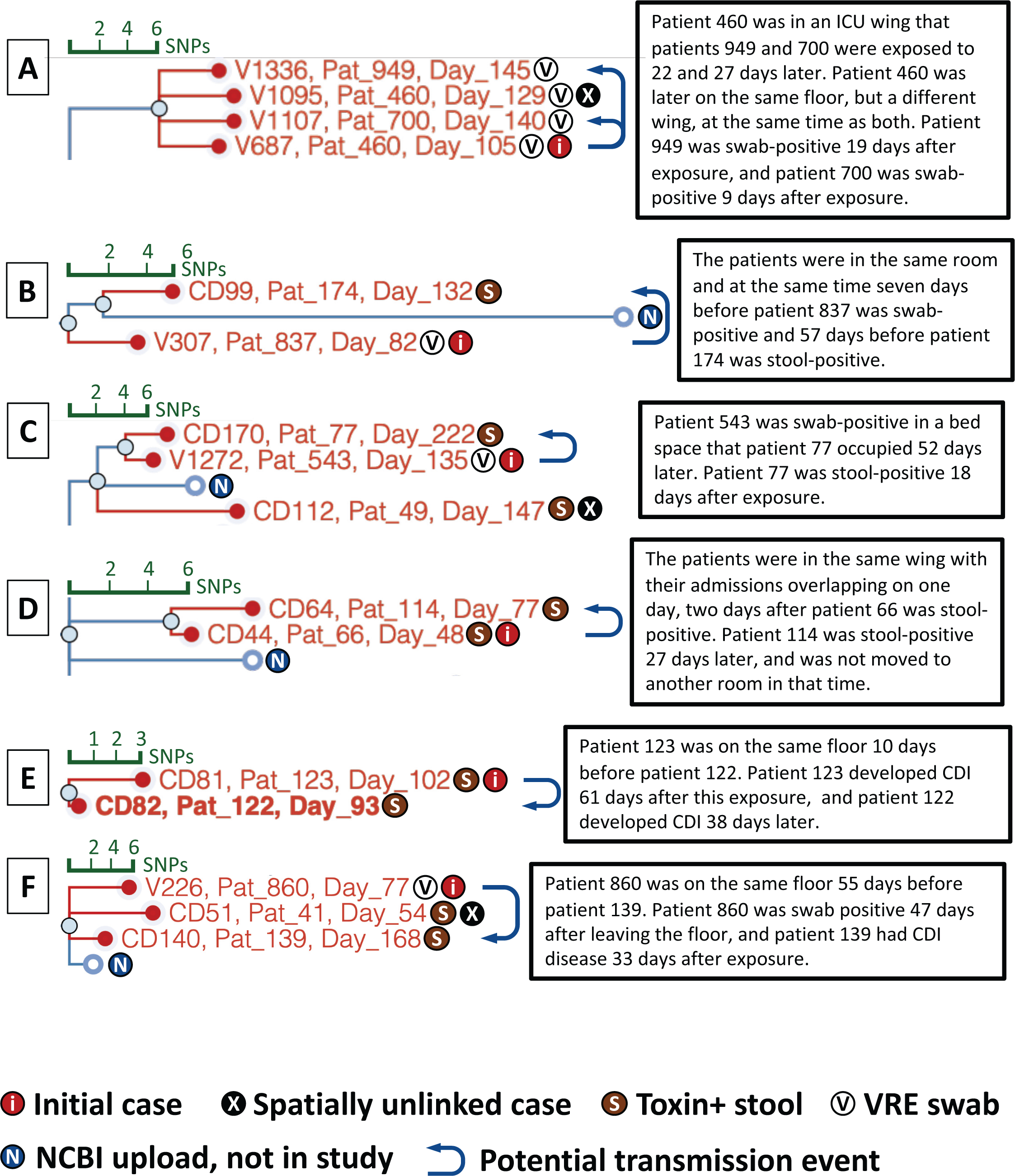
Cases with genetic and spatiotemporal evidence for nosocomial transmission. Leaves on the tree are labeled with isolate alias, <u>pat</u>ient number, and <u>day</u> of study the sample was collected after the beginning of stool collection. Images are taken directly from the NCBI Pathogen Detection Isolates Browser with user uploaded data.

Figure 6A shows an example analysis for asymptomatic transmission in genomically-identified strain clusters. The first patient, patient 460, produced swab isolate V687 on day 105, 2 days after their ICU admission. This patient remained colonized 24 days later during a second hospital admission to a different floor. Subsequent patients 949 and 700 had contact with the same ICU bed space and a room in the same wing, respectively, several weeks after this first case. Both patients tested positive for *C. difficile* by VRE-swab after this exposure. Patient 949 had an initial swab negative for *C. difficile* on day 126 before a second swab from day 145 produced isolate V1336, which occurred after a transfer to another room in another ICU.

Patients 460 and 949 both tested negative for *C. difficile* by stool EIA. While these patients were showing symptoms that led to CDI testing, the absence of toxin clinically by EIA suggests that they may have been colonized at a low level and did not have active CDI. The three cases thus represent potential asymptomatic transmission events from an initial asymptomatic carrier.

A spatial-temporal link was identified in patients 174 and 837 (Figure 6B) who occupied the same room concurrently. Patient 837 had confirmed asymptomatic colonization seven days later, while patient 174 was diagnosed with CDI 57 days later with a clonally-related isolate.

Other potential asymptomatic transmission cases demonstrated longer periods between spatial overlaps, including up to 52 days apart (Figures 6C-F, Supplemental Figure 2, 10).

## Discussion

We present the first detailed genomic and epidemiologic analyses of asymptomatic *C. difficile* carriage in ICU patients. The relative risk for developing active infection from asymptomatic carriage of a toxigenic strain was 9.32. Carriage was also associated with increased hospital lengths of stay and re-admissions during the study period, as well as 67% increased mortality in carriers when compared to mortality rates in non-colonized ICU patients. While the increased mortality seen in asymptomatic carriers represents a significant association, it is not a definitively causal relationship with *C. difficile* carriage, relative to other clinical factors, including that asymptomatic carriers had higher overall periods of exposure to the healthcare system. However, the potential for asymptomatic carriage to cause sub-clinical disease in patients with underlying co-morbidities raises a critical question on benefits of ICU screening for *C. difficile* to identify carriers, not only to reduce reservoirs for transmission, but to also reduce longer-term co-morbidities and mortality in carriers. Screening of vulnerable patient populations for *C. difficile* carriage also has potential to inform use of antibiotics and other clinical interventions to reduce risks for CDI in carriers [34].

Integrated genomic and epidemiologic analyses identified multiple potential transmission events from CDI patients and asymptomatic carriers to naïve patients. Analyses identified a 258-day window in which patient spatial overlaps were significantly associated with potential clonal transmission.

Asymptomatic carriers followed longitudinally carried the same strain, sometimes on the order of months [35,36]. Furthermore, asymptomatically carried strains caused active CDI in four of the five cases identified during the study period. In cases of extended periods between identification of asymptomatic carriage and CDI, we note that our findings cannot rule-out infection from direct carriage versus strain re-introduction from spores that persist in the patient’s environment [37].

We validated a culture-based method for *C. difficile* screening, leveraging samples from an existing VRE surveillance program. For centers with anaerobic culturing capabilities, the majority of cultures are negative and can be reported within 24 hours, offering a more cost-effective screening option over molecular methods [13,21,22]. As 30% of asymptomatic carriers were colonized with non-toxigenic strains, a finding that did not elevate risks for CDI, confirmation of toxigenicity by ELISA or PCR is warranted. Hospital-onset CDI costs an average of >$34,000 per patient. Prevention of even a subset of cases can bring significant savings in addition to improving patient outcomes [38]. Interventions for asymptomatic carriers could further reduce CDI incidence. A study placing *C. difficile-*colonized patients on contact precautions saw significant reductions in CDI [15]. As 5.1% of ICU-admitted patients were found to carry a toxigenic strain of *C. difficile*, the number of patients put on contact precautions would increase, potentially introducing burdens on clinical infrastructure. Informed by local rates of *C. difficile* carriage and CDI, healthcare facilities can assess the utility of screening by incorporating logistical and economic costs, as well as clinical actions to take upon identifying asymptomatic carriage [39].

Our epidemiologic analyses used a SNP cutoff of up to 17 SNPs to flag potential clonal clusters, a cutoff defined from analyses of longitudinal isolates from the same patient. Incorporation of strain genomic and patient spatio-temporal information identified potential *C. difficile* transmission events [40–42]. Analyses also validated use of publicly available SNP calling tools for *C. difficile* in the NCBI Pathogen Detection Isolates Browser [43,44]. Notably, SNP cluster PDS000036517 appeared in two healthcare institutions in the northeast and may represent a more prevalent cluster of *C. difficile*. As more institutions contribute *C. difficile* genomic data higher resolution analyses may be undertaken, particularly given the wide-spread nature of CDI across healthcare institutions.

Asymptomatic carriers of *C. difficile* provide a significant and hidden pathogen reservoir that can have negative adverse effects for carriers, other patients, and healthcare workers. We demonstrate constructive use of existing hospital surveillance programs and nationally available genomic tools and resources to support *C. difficile* surveillance within an ICU setting. Leveraging this model, institutions can make informed decisions regarding the utility of screening, and constructive actions to reduce CDI incidence.

## Data Availability

The genomic datasets have been deposited into the NCBI Short Read Archive and are available in the NCBI Pathogen Detection Browser at https://ncbi.nlm.nih.gov/pathogens/isolates
The C. difficile phylogeny created in this manuscript is available at https://itol.embl.de/tree/17022320725465491568233605

https://ncbi.nlm.nih.gov/pathogens/isolates

## Notes

### Funding

Funding was provided by the Hatch Family Foundation, the BWH Precision Medicine Institute, and NIDDK grant P30-DK034854. The work of Jay Worley was supported by the Intramural Research Program of the National Library of Medicine, National Institutes of Health.

### Conflicts of interest

No conflicts of interest are reported by the authors.

---

**Supplemental Table 1:** Patient characteristics by sample types.

|  | VRE-swab only | Stool only | Both |
| --- | --- | --- | --- |
| **Total number** | 1869 | 150 | 28 |
| **Age** | n=1749 | n=135 | n=27 |
| **Avg. age (std)** | 62.4 (16.0) | 60.7 (15.3) | 57.9 (15.9) |
| **Gender** | n=1749 | n=135 | n=27 |
| **% Female (num)** | 42.4 (n=755) | 51.9 (n=70) | 37.0 (n=10) |
| **% Male (num)** | 57.6 (n=1026) | 48.1 (n=65) | 63.0 (n=17) |
| **Ethnicity** | n=1716 | n=133 | n=27 |
| **% White (num)** | 82.2 (n=1411) | 88.0 (n=117) | 85.2 (n=23) |
| **% African American or Black (num)** | 8.9 (n=153) | 6.8 (n=9) | 3.7 (n=1) |
| **% Other** | 3.5 (n=60) | 0.8 (n=1) | 7.4 (n=2) |
| **% Asian (num)** | 2.7 (n=47) | 2.3 (n=3) | 0.0 (n=0) |
| **% Latino (num)** | 2.3 (n=40) | 2.3 (n=3) | 0.0 (n=0) |
| **% American Indian or Alaska Native (num)** | 0.2 (n=4) | 0.0 (n=0) | 3.7 (n=1) |
| **% Native Hawaiian or Pacific Islander (num)** | 0.0 (n=1) | 0.0 (n=0) | 0.0 (n=0) |
| **Hospital Stay during study** | n=1832 | n=144 | n=28 |
| **Avg. days inpatient (std)** | 14.7 (14.9) | 14.1 (16.3) | 38.8 (27.7)<sup>†</sup> |
| **Avg. number of inpatient admissions\* (std)** | 1.3 (0.8) | 1.4 (1.5) | 2.0 (1.1)<sup>†</sup> |
| **Avg. inpatient admission length (std)** | 12.4 (16.5)<br>(n=1799) | 13.8 (11.8) (n=91) | 26.3 (22.8) (n=27)<sup>†</sup> |
| **Outpatient days (std)** | 5.0 (7.4) | 5.3 (7.4) | 5.4 (9.1) |

std = standard deviation
num = Number reporting
\*Periods requiring official discharge due to inpatient status P
<sup>†</sup>p < 0.05 from both other categories using Mann-Whitney U after Kruskal-Wallis p < 0.01 across all groups. Only
tested for Hospital Stay statistics

**Supplemental Figure 1:**
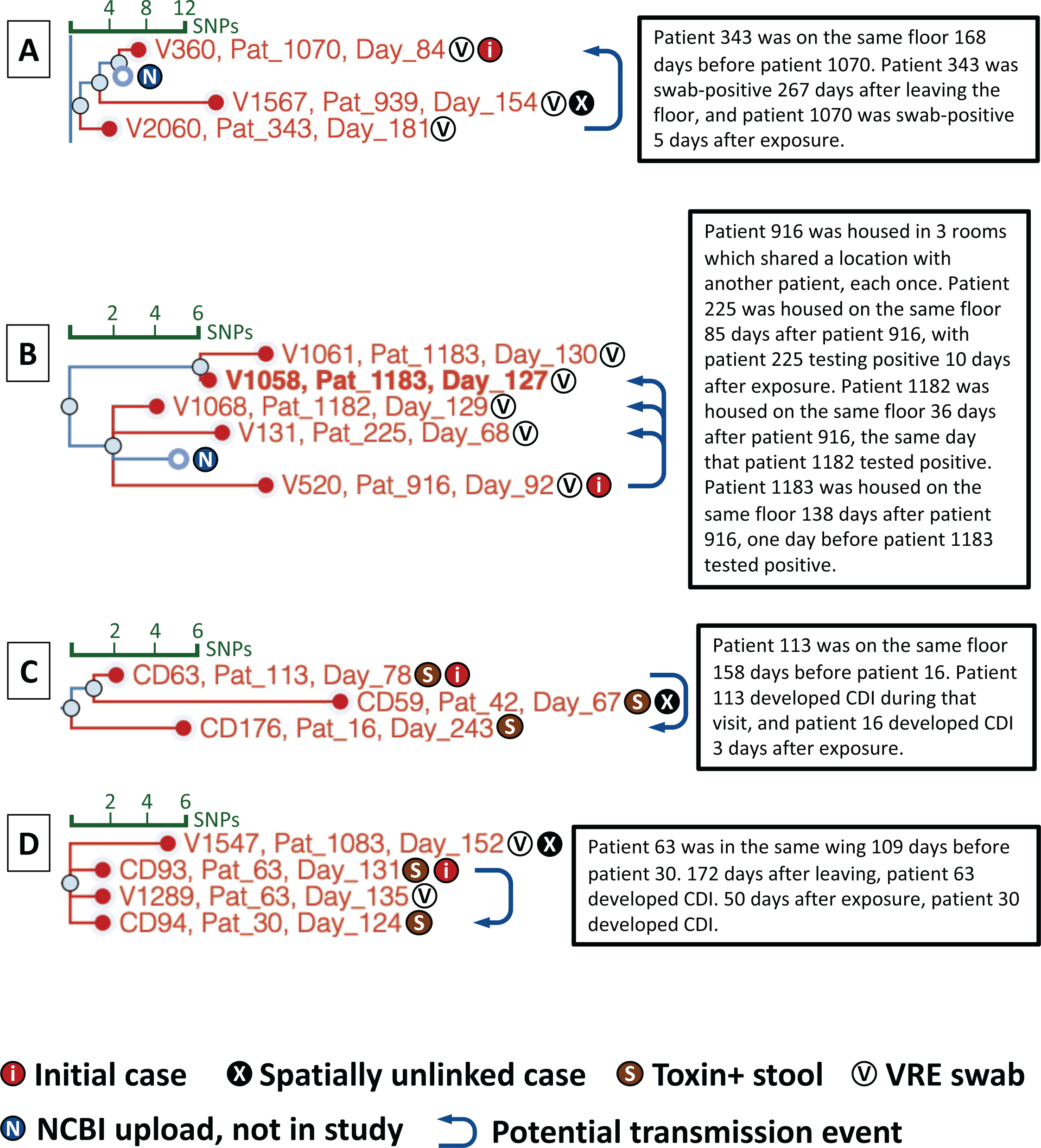
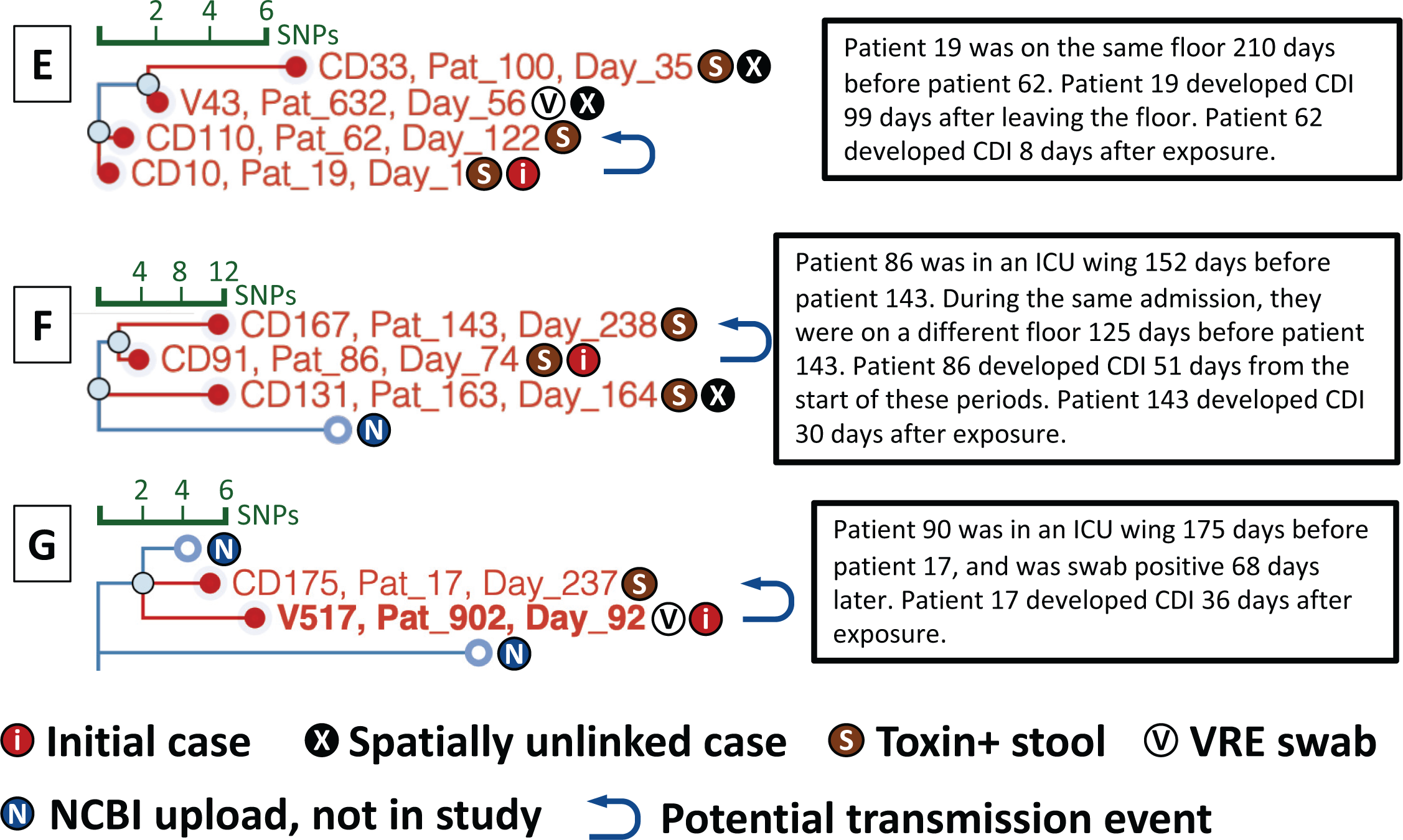
Additional cases with genetic and spatiotemporal evidence for nosocomial transmission. Leaves on the tree are labeled with isolate alias, <u>pat</u>ient number, and <u>day</u> of study the sample was collected after the beginning of stool collection. Images are taken directly from the NCBI Pathogen Detection Isolates Browser with user uploaded data.

## Notes

### Competing Interest Statement

The authors have declared no competing interest.

